# White Matter Microstructural Alterations and Symptom Correlates in Functional Motor Disorder

**DOI:** 10.64898/2026.08.25.26361322

**Authors:** Christiana Westlin, Cristina Bleier, Andrew J. Guthrie, Sara A. Finkelstein, Julie Maggio, Jessica Ranford, Julie MacLean, Ellen Godena, Daniel Millstein, Jennifer Freeburn, Caitlin Adams, Christopher D. Stephen, Marek Kubicki, Ibai Diez, David L. Perez

**Author notes:** **Corresponding authors**: Christiana Westlin, PhD; Massachusetts General Hospital, 55 Fruit Street, Boston, MA, USA, 02114;, David L. Perez, MD, MMSc; Massachusetts General Hospital, 55 Fruit Street, Boston, MA, USA, 02114. Denotes equally contributing co-senior authors (Ibai Diez, David L. Perez).

## Abstract

**Background:** Neuroimaging studies implicate network alterations in functional motor disorder (FND-motor), yet white matter remains poorly characterized.

**Objectives:** To characterize white matter microstructure in FND-motor relative to healthy (HCs) and psychiatric (PCs) controls and examine symptom associations.

**Methods:** Fifty individuals with FND-motor, 50 age- and sex-matched HCs, and 50 PCs matched on age, sex, depression, anxiety, and post-traumatic stress disorder severity underwent multi-shell diffusion MRI. Voxel-based analyses examined whole-brain white matter using diffusion tensor imaging (fractional anisotropy [FA], mean diffusivity [MD]) and neurite orientation dispersion and density imaging (NODDI) (neurite density index [NDI], orientation dispersion index, and free water fraction [FWF]) metrics. Cross-metric convergence was characterized using atlas-based tract overlap analyses and probabilistic tractography. Associations with FND symptoms and transdiagnostic physical symptoms were also evaluated.

**Results:** Compared with HCs, FND-motor showed higher FA/NDI and lower MD/FWF, predominantly in the middle cerebellar peduncle. Compared with PCs, differences were limited to lower MD/FWF, involving the corpus callosum, middle cerebellar peduncle, and left inferior longitudinal fasciculus. Greater FND symptom severity was associated with a lower FA/NDI and higher MD/FWF in the corpus callosum and right-lateralized association and projection pathways, whereas greater transdiagnostic physical symptom burden across FND-motor and PCs was associated with higher FA and lower MD/FWF in the middle cerebellar peduncle.

**Conclusions:** This study provides a comprehensive multi-metric diffusion-weighted characterization of white matter microstructure in FND-motor relative to both HCs and PCs – highlighting cortico-cerebellar connections via the middle cerebellar peduncle as distinct in FND-motor and associated transdiagnostically with physical symptom burden.

## INTRODUCTION

Functional neurological disorder (FND) is a common and potentially disabling neuropsychiatric condition characterized by motor, sensory, and cognitive symptoms [1]. The motor variant of FND (FND-motor), including functional tremor, limb weakness, gait difficulties, jerky movements, and dystonia, is a prevalent subtype associated with substantial costs and reduced quality of life [2–4]. Brain imaging studies in FND-motor have identified alterations in functional activation, connectivity, and grey matter structure in regions of the somatomotor, salience/ventral attention, and default mode networks, among others, yet comparatively less is known about white matter microstructure [5,6].

Diffusion-weighted MRI studies have predominantly focused on mixed FND cohorts, functional seizures, or functional dystonia [7–13]. These studies have reported alterations in several white matter tracts, including the corpus callosum, corticospinal tract, uncinate fasciculus, stria terminalis/fornix, and cingulum bundle. However, findings have been heterogeneous, with some studies reporting widespread abnormalities and others identifying more limited or phenotype-specific alterations. Additionally, most studies have relied on conventional diffusion tensor imaging (DTI) metrics, which provide sensitive but nonspecific markers of white matter organization. Attenuation of some past findings after accounting for depression and anxiety severity also suggests that some alterations may be influenced by affective symptoms [13]. Moreover, most studies have compared patients to healthy controls (HCs), limiting the ability to determine whether white matter alterations are specific to FND or reflect features shared with common psychiatric comorbidities.

Multi-shell diffusion MRI allows white matter to be characterized across complementary diffusion metrics. Conventional DTI metrics, including fractional anisotropy (FA) and mean diffusivity (MD), provide sensitive but biologically nonspecific indices of directional and average water molecule diffusion, respectively [14]. In contrast, neurite orientation dispersion and density imaging (NODDI) offers greater biological specificity by estimating the packing density of neurites (neurite density index, NDI), the orientation coherence of neurites (orientation dispersion index, ODI), and the signal within a voxel that comes from freely diffusing water (free water fraction, FWF; [15]). NODDI has previously been applied to functional seizures [9–11], but has not, to our knowledge, been used in FND-motor.

Here, we investigated whole-brain white matter microstructure in 50 individuals with FND-motor compared to 50 age- and sex-matched HCs and 50 psychiatric controls (PCs) matched at the group-level on age, sex, depression, anxiety, and post-traumatic stress disorder (PTSD) severity. We combined DTI and NODDI metrics, identified areas demonstrating convergent cross-metric differences in FND-motor, and anatomically characterized these findings using atlas-based tract overlap analyses and probabilistic tractography. We also examined relationships between white matter microstructure and both FND symptom severity and transdiagnostic physical symptom burden. By including PCs with comparable affective symptom burden but without FND, we sought to distinguish alterations more closely associated with FND-motor from those that may reflect shared psychiatric comorbidity. Together, these analyses aimed to delineate white matter microstructural alterations associated with FND-motor and to examine white matter relationships with clinically-relevant symptom dimensions, providing insights into the pathophysiology of the disorder.

## METHODS

### Participants

Fifty participants with FND-motor (42 females, 8 males; mean age=39.9±13.2; average illness duration 2.8±3.0 years (range=0.3-16.1 years) were prospectively recruited from the Massachusetts General Hospital between January 2019 and December 2025 (**Table 1**). FND diagnoses were based on positive examination signs [2]. Exclusion criteria were major neurological comorbidities (e.g., Parkinson’s disease), known brain MRI abnormalities, poorly controlled medical problems with central nervous system consequences, active illicit substance dependence, known psychosis, and/or active suicidality. FND-motor is defined as the motor variant of FND, inclusive of functional movement disorder and functional limb weakness; FND-motor cohort phenotypic breakdown was: tremor=24, weakness=22; gait=20; tics/jerks/spasms=8; dystonia=4 [phenotypes were not mutually exclusive; **Supplementary Figure 1, Supplementary Table 1**]; 10 also met criteria for functional seizures (7 documented, 1 clinically-established, 2 probable). FND-motor individuals also had a history of depression (*n*=31), anxiety (*n*=35), and/or PTSD (*n=*25); 32 (64%) had more than one of these diagnoses. Data from this cohort have been published in structural (grey matter) and functional MRI studies [16–18]; diffusion-weighted MRI data has not been previously published.

**Table 1.**
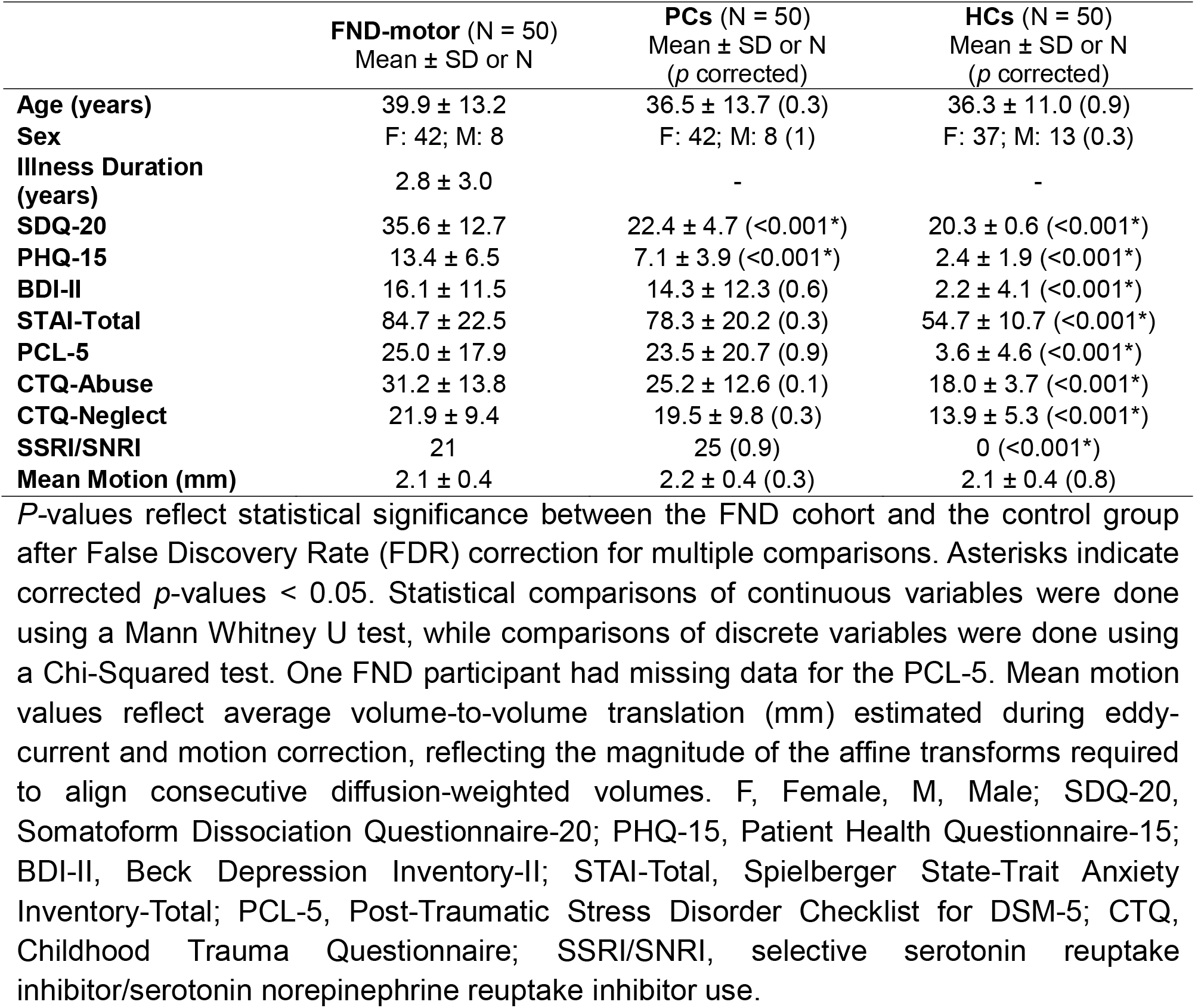
Demographic and psychometric characteristics of the functional motor disorder (FND-motor), psychiatric control (PC) and healthy control (HC) samples.

Fifty PCs (42 females, 8 males; mean age=36.5±13.7) were recruited from the community and were matched to the FND cohort on age, sex, depression, anxiety, and PTSD-severity, as assessed during neuropsychiatric characterization (see below). PCs had a history of depression (*n*=43), anxiety (*n*=40), and/or PTSD (*n=*20); 40 (80%) had more than one diagnosis. Psychotropic medication use was permitted in both the FND-motor and PC cohorts; SSRI/SNRI use was included as a covariate of non-interest in all neuroimaging analyses. Exclusion criteria were the same as for the FND-motor cohort, with the added exclusion of FND or somatic symptom disorder diagnoses (**Supplementary Table 2**).

Fifty age- and sex-matched HCs (37 females, 13 males; mean age=36.3±11.0) were recruited from the community. HCs had no history of psychiatric disorders, and none were on psychotropic medications. Exclusion criteria otherwise matched those of the FND-motor and PC cohorts. All subjects provided informed consent, and the Mass General Brigham Institutional Review Board approved this study.

### Neuropsychiatric Characterization

Participants completed a Structured Clinical Interview for Diagnostic and Statistical Manual Disorders (SCID-I), as well as questionnaires including the Beck Depression Inventory-II (BDI-II; [19]), Spielberger State-Trait Anxiety Inventory (STAI; [20]), PTSD Checklist-5 (PCL-5; [21]), and Childhood Trauma Questionnaire (CTQ; [22]). FND symptom severity was measured using the Somatoform Dissociation Questionnaire-20 (SDQ-20; [23]), a 20-item measure of the extent to which FND symptoms (e.g., paralysis, body stiffening) were experienced over the past year. Transdiagnostic physical symptom severity was measured using the Patient Health Questionnaire-15 (PHQ-15; [24]), a 15-item measure of how bothersome physical symptoms (e.g., pain, fatigue) were over the past four weeks.

### MRI Acquisition and Preprocessing

Participants were scanned on the same Simens Tim Trio 3T MRI scanner using a 12-channel phased-array head coil, acquiring a high-resolution T1-weighted magnetization-prepared rapid gradient echo (MP-RAGE) sequence and a multi-shell, diffusion-weighted, spin-echo echo-planar imaging (EPI) sequence. For DTI, participant-level voxel-wise FA and MD maps were extracted. For NODDI, participant-level voxel-wise NDI, ODI, and FWF maps were generated. See Supplementary Methods for sequence and pre-processing details.

### Voxel-Based Analyses

Voxel-based analyses (VBA) were used to characterize whole-brain white matter microstructural alterations (as previously described [12]). For each diffusion metric, individual maps were transformed to MNI152 standard space and smoothed with an isotropic Gaussian kernel (sigma=2mm). Analyses were restricted to a white matter mask, derived from the Human Connectome Project [25]. Cluster-wise correction for multiple comparisons was performed using a Monte Carlo simulation with 10,000 iterations to estimate the probability of false positive clusters at *p*<0.05.

### Between-group comparisons

Voxel-wise general linear models (GLMs) were used to compare FND-motor to HCs and PCs. Primary models included age, sex, head motion (average volume-to-volume translation), selective serotonin reuptake inhibitor/serotonin-norepinephrine reuptake inhibitor (SSRI/SNRI) use, and co-occurring functional seizures (yes/no) as covariates of non-interest. *Post-hoc* models also adjusted for (i) BDI-II, STAI-total, PCL-5 scores, and (ii) CTQ-abuse and CTQ-neglect scores. Intersection maps were computed to examine significant voxels across primary and *post-hoc* adjustments. Additional analyses repeated the primary comparisons in females only [26,27].

### Symptom associations

Within the FND-motor cohort, voxel-wise GLMs evaluated associations between diffusion metrics (whole brain, white matter restricted) and SDQ-20 scores. Across FND-motor and PCs, voxel-wise GLMs evaluated associations between diffusion metrics and PHQ-15 scores. Covariate adjustments matched those described above for between-group comparisons.

### Cross-metric convergence

Convergence maps were generated to identify white matter regions demonstrating consistent findings across DTI and NODDI metrics. Convergent voxels were used as seed regions for probabilistic tractography and tract overlap analyses.

### Anatomical Characterization of Convergent Findings

To characterize convergent white matter findings, we used two complementary approaches. First, convergent voxels were quantified according to their overlap with white matter tracts from the HCP-1065 Population-Averaged Tractography Atlas [28]. Second, convergent voxels were used as seed regions for probabilistic tractography using diffusion-weighted MRI data from the 50 HCs, and the resulting projection maps were similarly quantified using atlas-based tract overlap. Tracts with >5% overlap are reported. Whereas atlas overlap localized the significant voxels themselves, atlas overlap of the probabilistic tractography maps characterized the broader pathways traversing those voxels. See **Supplementary Methods** for full probabilistic tractography and tract overlap procedures.

## RESULTS

### Demographic and Psychometric Comparisons

There were no age, sex, or mean head motion differences between FND-motor, PCs, and HCs. Compared with PCs, FND-motor had higher SDQ-20 and PHQ-15 scores, but did not differ on BDI-II, STAI-total, PCL-5, or CTQ-abuse or neglect subscales; there were also no differences in SSRI/SNRI use between FND-motor and PCs. Compared to HCs, FND-motor had higher scores on all psychometrics (**Table 1**).

### Between-Group Comparisons

### FND-motor vs. HCs

VBA identified white matter differences between FND-motor and HCs across all five metrics (**Figure 1**; **Supplementary Figure 2**). The most convergent cross-metric pattern was characterized by higher FA and NDI, and lower MD and FWF in FND-motor relative to HCs (**Figure 1** **A/B**). Based on overlap with the HCP1065 tractography atlas, convergent voxels localized primarily to the middle cerebellar peduncle (72.4%), as well as left corticospinal (6.8%), parietal corticopontine (5.3%), and corticobulbar (5.1%) tracts (**Table 2**). Probabilistic tractography seeded from convergent voxels demonstrated a similar pattern, with projections predominantly traversing the middle cerebellar peduncle (73.2% of weighted overlap), followed by left corticospinal (8.1%) and parietal corticopontine (6.7%) tracts (**Figure 1C**; **Supplementary Table 3**). Mean values extracted from convergent voxels were consistent with the voxel-wise findings, with FND-motor showing higher FA/NDI, and lower MD/FWF than HCs (**Figure 1B**); PCs, shown for visualization only, exhibited intermediate mean values across the four metrics (see **Supplementary Figure 4A** for plots by FND-motor phenotype). Findings predominantly remained significant when adjusting for (i) depression, anxiety, and PTSD scores, (ii) childhood trauma, and in female-only analyses (**Supplementary Figure 2**), with the exception of FA, which attenuated after adjustment for depression, anxiety, and PTSD scores. See **Supplementary Figure 2** for additional observations.

**Figure 1.**
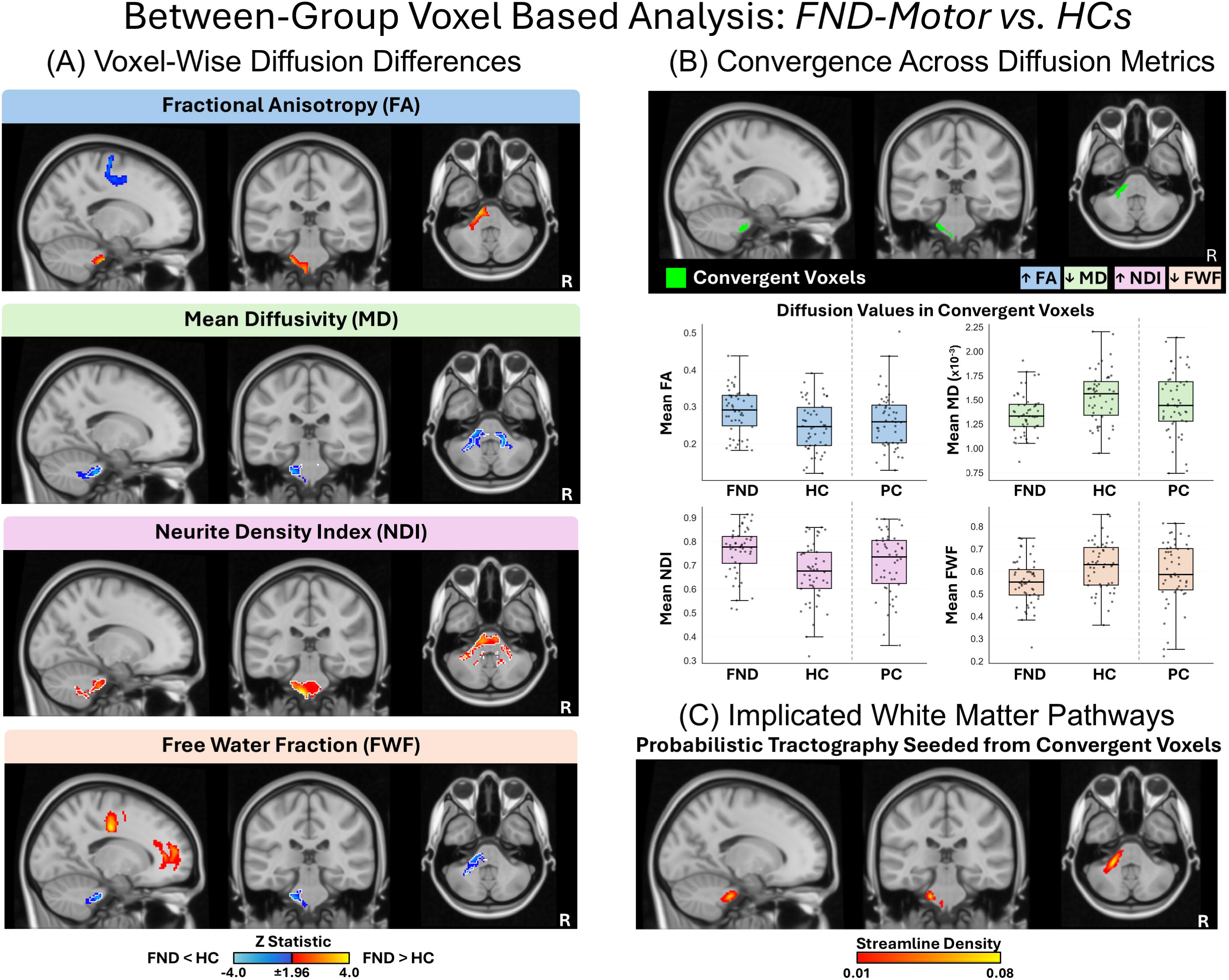
White matter microstructural alterations in functional motor disorder (FND-motor) compared with healthy controls (HCs). **(A)** Results from voxel-based analyses comparing individuals with FND-motor (N=50) and HCs (N=50). Between-group differences are shown for fractional anisotropy (FA), mean diffusivity (MD), neurite density index (NDI), and free water fraction (FWF). Colors reflect the z-statistic computed from the primary voxel-wise general linear model adjusting for age, sex, head motion, SSRI/SNRI use, and co-occurring functional seizures (z > 1.96, *p* < 0.05 cluster-corrected for multiple comparisons). Warm colors indicate higher diffusion metric values in FND-motor vs. HCs, whereas cool colors indicate lower values. White outlines reflect regions that held across all *post-hoc* corrections for (i) BDI-II, STAI-total, and PCL-5 scores, and (ii) CTQ-abuse and CTQ-neglect scores. Statistical maps for *post-hoc* comparison are visualized in **Supplementary** Figure 2. **(B)** Cross-metric convergence map showing the intersection of significant voxel-wise findings across FA, MD, NDI and FWF. Convergent voxels exhibited higher FA, lower MD, higher NDI, and lower FWF in FND-motor vs. HCs. Boxplots show mean diffusion values extracted from the convergent voxels for FND-motor, HCs, and psychiatric controls (PCs), where each dot reflects a single participant’s mean value. PCs are displayed for visualization only and were not included in the statistical comparison. **(C)** Probabilistic tractography seeded from the convergent voxels, illustrating the likely implicated white matter pathways. Tractography was conducted across the HC cohort (N=50). Colors indicate the relative density of probabilistic streamlines traversing each voxel, averaged across HCs, with brighter yellow indicating a greater density of streamlines. Atlas-based tract overlap analyses identifying the white matter tracts represented by the convergent voxels and associated probabilistic tractography are summarized in **Table 2** and **Supplementary Table 3**. SSRI/SNRI, selective serotonin reuptake inhibitor/serotonin-norepinephrine reuptake inhibitor.

**Table 2.** Overlap between convergent white matter voxels identified in voxel-based analyses and HCP-1065 white matter tracts.

| **Between-Group Voxel-Based Analyses** |  |  |  |
| --- | --- | --- | --- |
|  | **Overlapping HCP-1065 Atlas Tract** | **Number of Overlapping Voxels** | **Percent of Total Overlap** |
| **FND-motor vs. HCs**<br>VBA Convergent Voxels<br>Across FA, MD, NDI,<br>FWF | Middle Cerebellar Peduncle | 343 | 72.4% |
|  | Left Corticospinal Tract | 32 | 6.8% |
|  | Left Parietal Corticopontine Tract | 25 | 5.3% |
|  | Left Corticobulbar Tract | 24 | 5.1% |
| **FND-motor vs. PCs**<br>VBA Convergent Voxels<br>Across MD, FWF | Corpus Callosum | 955 | 26.9% |
|  | Middle Cerebellar Peduncle | 377 | 10.6% |
|  | Left Inferior Longitudinal Fasciculus | 358 | 10.1% |
|  | Left Medial Lemniscus | 298 | 8.4% |
|  | Left Optic Radiation | 276 | 7.8% |
|  | Anterior Commissure | 269 | 7.6% |
|  | Right Medial Lemniscus | 266 | 7.5% |
| **Associations With Clinical Features** |  |  |  |
|  | **Overlapping HCP-1065 Atlas Tract** | **Number of Overlapping Voxels** | **Percent of Total Overlap** |
| **Associations with SDQ-20**<br>VBA Convergent Voxels<br>Across FA, MD, NDI | Corpus Callosum | 790 | 28.6% |
|  | Right Parietal Corticopontine Tract | 537 | 19.4% |
|  | Right Superior Thalamic Radiation | 474 | 17.1% |
|  | Right Superior Corticostriatal Tract | 335 | 12.1% |
|  | Right Medial Lemniscus | 313 | 11.3% |
| **Associations with SDQ-20**<br>VBA Convergent Voxels<br>Across FA, MD, FWF | Corpus Callosum | 773 | 14.8% |
|  | Right Superior Longitudinal Fasciculus 2 | 599 | 11.4% |
|  | Right Corticospinal Tract | 571 | 10.9% |
|  | Right Superior Corticostriatal Tract | 518 | 9.9% |
|  | Right Parietal Corticopontine Tract | 517 | 9.9% |
|  | Right Superior Thalamic Radiation | 489 | 9.3% |
|  | Right Frontal Corticostriatal Tract | 409 | 7.8% |
| **Associations with PHQ-15**<br>VBA Convergent Voxels<br>Across FA, MD, FWF | Middle Cerebellar Peduncle | 374 | 87.8% |
|  | Left Cerebellum WM | 39 | 9.2% |

Overlapping voxels indicates the number of convergent voxels located within each tract ROI. Percent of total overlap indicates the proportion of all overlapping voxels across HCP-1065 tract ROIs accounted for by a given tract. Results are shown for tracts with >5 percent of total overlap. HCs, healthy controls; PC, psychiatric controls; VBA, voxel-based analysis; FA, fractional anisotropy; MD, mean diffusivity; NDI, neurite density index; FWF, free water fraction; WM, white matter.

### FND-motor vs. PCs

VBA identified white matter microstructural differences between FND-motor and PCs across MD and FWF (**Figure 2A; Supplementary Figure 3**). Across these two metrics, a convergent pattern was characterized by lower MD and FWF in FND-motor vs. PCs (**Figure 2B**). Based on overlap with the HCP-1065 tractography atlas, convergent voxels localized to the posterior corpus callosum (26.9%), as well as middle cerebellar peduncle (10.6%), bilateral medial lemniscus (left:8.4%, right:7.5%), left optic radiation (7.8%), and anterior commissure (7.6%) (**Table 2**). Probabilistic tractography seeded from convergent voxels revealed projections predominantly traversing the posterior corpus callosum (31.3% of weighted overlap), left inferior longitudinal fasciculus (16.3%), anterior commissure (15.9%), left inferior fronto-occipital fasciculus (12.2%), and left optic radiation (11.9%) (**Figure 2C**; **Supplementary Table 3**). Mean values across convergent values demonstrated FND-motor exhibiting lower mean MD/FWF than PCs, with HCs, shown for visualization only, exhibiting intermediate mean values (**Figure 2B**; see **Supplementary Figure 4B** for plots by FND-motor phenotype). MD differences were most robust within cerebellar regions, surviving *post-hoc* and female-only analyses. In contrast, FWF differences within cortical white matter survived *post-hoc* analyses, particularly in voxels overlapping with the posterior corpus callosum and left inferior longitudinal fasciculus, whereas cerebellar FWF findings attenuated following *post-hoc* adjustments, and demonstrated a distinct spatial female-only pattern.

**Figure 2.**
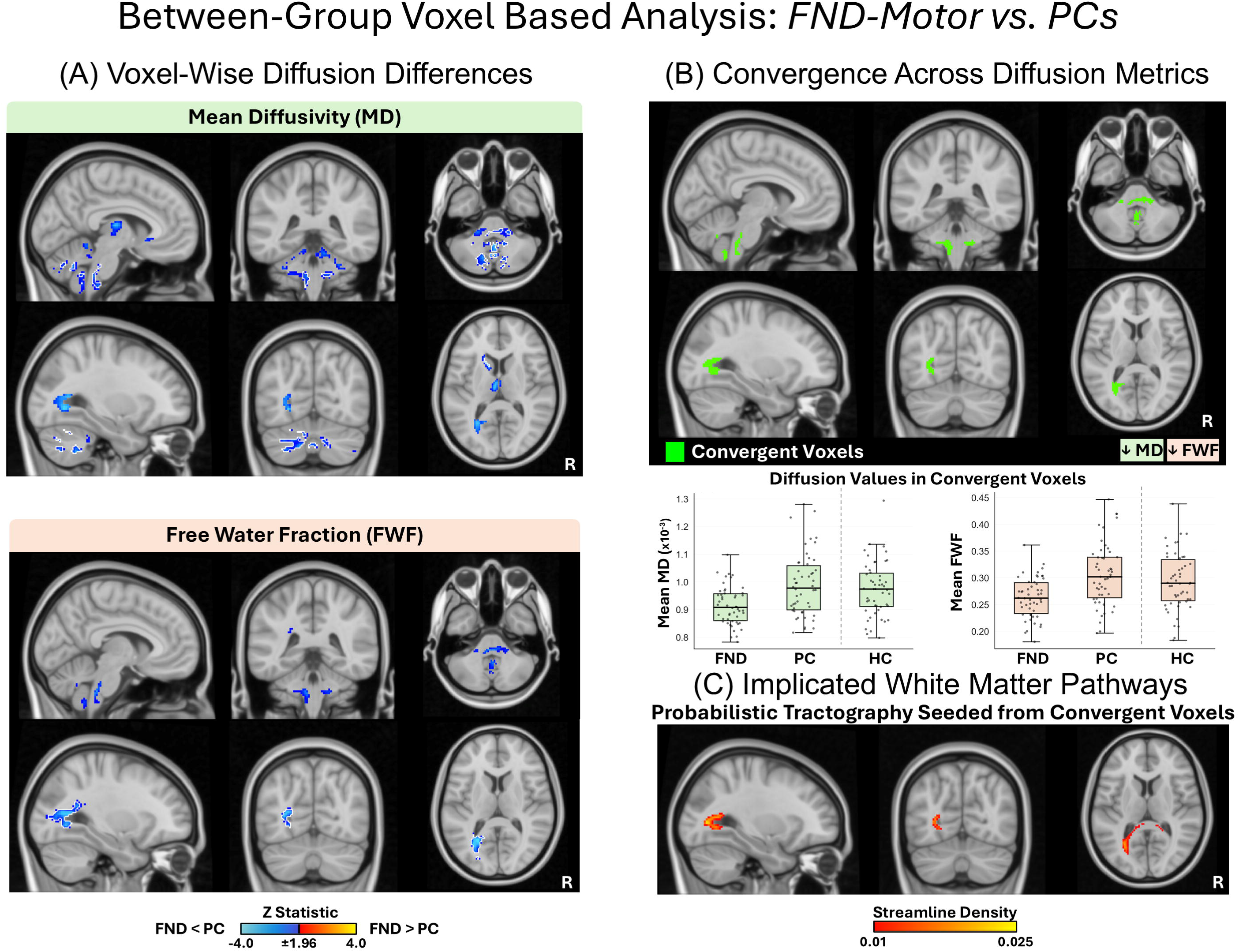
White matter microstructural alterations in functional motor disorder (FND-motor) compared with psychiatric controls (PCs). **(A)** Results from voxel-based analyses comparing individuals with FND-motor (N=50) and PCs (N=50). Between-group differences are shown for mean diffusivity (MD) and free water fraction (FWF). Colors reflect the z-statistic computed from the primary voxel-wise general linear model adjusting for age, sex, head motion, SSRI/SNRI use, and co-occurring functional seizures (z > 1.96, *p* < 0.05 cluster-corrected for multiple comparisons). Cool colors indicate lower diffusion metric values in FND-motor vs. PCs. White outlines reflect regions that held across all *post-hoc* corrections for (i) BDI-II, STAI-total, and PCL-5 scores, and (ii) CTQ-abuse and CTQ-neglect scores. Statistical maps for *post-hoc* comparison are visualized in **Supplementary** Figure 3. **(B)** Cross-metric convergence map showing the intersection of significant voxel-wise findings across MD and FWF. Convergent voxels exhibited lower MD and FWF in FND-motor relative to PCs. Boxplots show mean diffusion values extracted from the convergent voxels for FND-motor, PCs, and healthy controls (HCs), where each dot reflects a single participant’s mean value. HCs are displayed for visualization only and were not included in the statistical comparison. **(C)** Probabilistic tractography seeded from the convergent voxels, illustrating the likely implicated white matter pathways. Tractography was conducted across the HC cohort (N=50). Colors indicate the relative density of probabilistic streamlines traversing each voxel, averaged across HCs, with brighter yellow indicating a greater density of streamlines. Atlas-based tract overlap analyses identifying the white matter tracts represented by the convergent voxels and associated probabilistic tractography are summarized in **Table 2** and **Supplementary Table 3**. SSRI/SNRI, selective serotonin reuptake inhibitor/serotonin-norepinephrine reuptake inhibitor.

### Symptom Associations

#### SDQ-20

Within the FND-motor cohort, VBA identified associations between microstructure and SDQ-20 scores across FA, MD, NDI, and FWF (**Figure 3A**; **Supplementary Figure 5**). Two convergent patterns were identified, characterized by (i) lower FA and NDI, and higher MD, associated with higher SDQ-20 scores; and (ii) lower FA, and higher MD and FWF, associated with higher SDQ-20 scores (**Figure 3A**). Based on overlap with the HCP-1065 tractography atlas, convergent voxels across FA/MD/NDI overlapped primarily with the mid-posterior corpus callosum (28.6%), right parietal corticopontine (19.4%), right superior thalamic radiation (17.1%), right superior corticostriatal (12.1%), and right medial lemniscus (11.3%) tracts (**Table 2**). Probabilistic tractography seeded from these convergent voxels projected through the mid-posterior corpus callosum (26.7% of weighted overlap), and right parietal corticopontine (20.4%), superior thalamic radiation (14.7%), medial lemniscus (12.1%), and superior corticostriatal (11.7%) tracts (**Supplementary Figure 7A; Supplementary Table 3**). The second convergence map (FA/MD/FWF) demonstrated similar anatomical overlap. Based on overlap with the HCP-1065 tractography atlas, convergent voxels overlapped with the central corpus callosum (14.8%), and right superior longitudinal fasciculus II (11.4%), corticospinal (10.9%), superior corticostriatal (9.9%), parietal corticopontine (9.9%), superior thalamic radiation (9.3%), and frontal corticostriatal (7.8%) tracts (**Table 2**). Probabilistic tractography seeded from these convergent voxels projected predominantly through the right parietal corticopontine (13.2% of weighted overlap), central corpus callosum (10.7%), and right superior thalamic radiation (9.5%), superior corticostriatal (9.4%), frontal corticopontine (8.9%), frontal corticostriatal (8.0%), superior longitudinal fasciculus II (6.6%), dentatorubrothalamic (6.2%), and medial lemniscus (5.0%) tracts (**Supplementary Figure 7B; Supplementary Table 3**). Findings across both convergent patterns remained largely significant following *post-hoc* adjustments and were present in female-only analyses, although NDI findings attenuated following adjustment for depression, anxiety, and PTSD scores. Additional positive associations between FA and SDQ-20 within the middle cerebellar peduncle were also observed but did not demonstrate cross-metric convergence (**Supplementary Figure 5).**

**Figure 3.**
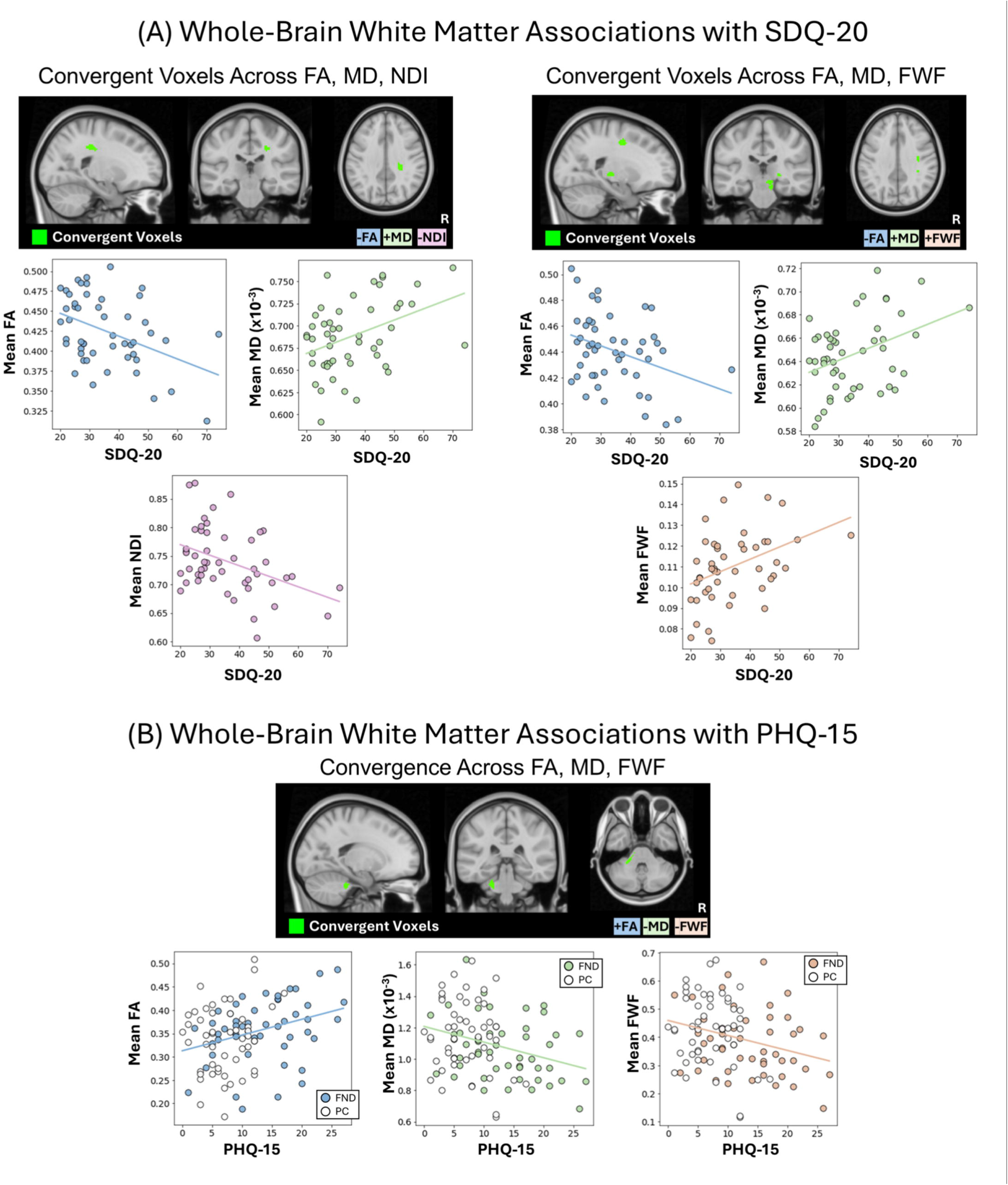
Whole-brain white matter associations with symptom severity. **(A)** Whole-brain white matter associations with SDQ-20 symptom severity. Cross-metric convergence map showing the intersection of significant voxel-wise associations between SDQ-20 scores and white matter microstructure in individuals with functional motor disorder (FND-motor; N=50). ***Left:*** Convergence across fractional anisotropy (FA), mean diffusivity (MD), and neurite density index (NDI), with positive associations for FA and NDI and negative associations for MD. ***Right:*** Convergence across FA, MD, and free water fraction (FWF), with positive associations for FA and negative associations for MD and FWF. Scatterplots show SDQ-20 scores plotted against mean diffusion values extracted from the convergent voxels (FA, MD, and NDI or FWF), where each dot represents a single participant. Regression lines are shown for visualization only. Voxel-wise association maps for each diffusion metric and corresponding *post-hoc* analyses are visualized in **Supplementary** Figure 5. Atlas-based tract overlap analyses identifying the white matter tracts represented by the convergent voxels are summarized in **Table 2**. Probabilistic tractography seeded from the convergent voxels, and corresponding atlas-based tract overlap analyses, are shown in **Supplementary** Figure 7 and **Supplementary Table 3**. (**B**) Whole-brain white matter associations with PHQ-15 symptom severity. Cross-metric convergence map showing the intersection of significant voxel-wise associations across fractional anisotropy (FA), mean diffusivity (MD), and free water fraction (FWF) in analyses relating white matter microstructure to individual differences in PHQ-15 scores in individuals with functional motor disorder (FND-motor; N=50) and psychiatric controls (PCs; N=50). Convergent voxels demonstrated positive associations for FA and negative associations for MD and FWF. Voxel-wise association maps for each diffusion metric and corresponding *post-hoc* analyses are visualized in **Supplementary** Figure 6. Atlas-based tract overlap analyses identifying tracts represented by the convergent voxels are summarized in **Table 2**. Probabilistic tractography seeded from the convergent voxels, and corresponding atlas-based tract overlap analyses, are shown in **Supplementary** Figure 7 and **Supplementary Table 3**. Scatterplots show PHQ-15 scores plotted against mean diffusion values extracted from the convergent voxels (left: FA; middle: MD; right: FWF), where each dot represents a single participant. Colored dots denote participants with FND-motor, and open dots denote PCs. Regression lines are shown for visualization only.

#### PHQ-15

Across FND-motor and PCs, VBA identified associations between all five diffusion metrics and PHQ-15 scores (**Figure 3B**; **Supplementary Figure 6**). The primary convergent pattern was characterized by higher FA, and lower MD and FWF, associated with higher PHQ-15 scores. Based on overlap with HCP-1065 tractography atlas, convergent voxels overlapped predominantly with the middle cerebellar peduncle (87.8%), as well as left cerebellar white matter (9.2%) (**Table 2**). Probabilistic tractography seeded from convergent voxels projected predominantly through the middle cerebellar peduncle (76.3%), with additional projections to left cerebellar white matter (13.4%) (**Supplementary Figure 7C**; **Supplementary Table 3**). Findings remained significant in *post-hoc* corrections for depression, anxiety, and PTSD severity, and were present in female-only analyses; when correcting *post-hoc* for childhood trauma, MD and FWF findings attenuated. See **Supplementary Figure 6** for additional spatially distinct and non-convergent observations.

## DISCUSSION

In this study, we examined white matter microstructure in FND-motor using DTI and NODDI, employing between-group voxel-based analyses against HCs and PCs, as well as analyses relating white matter microstructure to individual differences in FND symptom severity and transdiagnostic physical symptom burden. Compared with HCs, FND-motor exhibited convergent white matter differences in the middle cerebellar peduncle and cortico-ponto-cerebellar pathways, characterized by higher FA and NDI, and lower MD and FWF. Compared with PCs, FND-motor exhibited a partially distinct pattern involving the posterior corpus callosum, middle cerebellar peduncle, and inferior longitudinal fasciculus, characterized by lower MD and FWF. Within FND-motor, higher FND symptom severity was associated with lower FA and NDI, and higher MD and FWF across the mid-posterior and central corpus callosum and predominantly right-lateralized association and projection pathways, whereas greater transdiagnostic physical symptom burden across FND-motor and PCs was associated with higher FA and NDI, and lower MD and FWF, in the middle cerebellar peduncle. Across analyses, findings were generally robust to *post-hoc* adjustment for affective symptom severity and childhood trauma, although some findings attenuated following these adjustments. Together, these findings identify convergent and dissociable patterns of white matter microstructure across group comparisons and symptom associations.

In FND-motor vs. HC comparisons, convergent alterations within the middle cerebellar peduncle and broader cortico-ponto-cerebellar pathways were characterized by higher FA and NDI alongside lower MD and FWF, a pattern likely reflective of altered white matter microstructure in FND-motor. Findings largely remained significant in *post-hoc* corrections for affective symptoms and trauma burden, although FA findings attenuated following correction for depression, anxiety, and PTSD severity, suggesting that these factors may play a role in some of the observed differences. Probabilistic tractography similarly implicated the middle cerebellar peduncle as the predominant tract, followed by corticospinal and corticopontine tracts, providing additional support for altered cortico-cerebellar motor pathways in FND-motor. Middle cerebellar peduncle white matter differences are noteworthy given its role in cortico-cerebellar communication; as the principal afferent pathway to the cerebellum, it carries fibers from the pontine nuclei to the cerebellum, relaying information from widespread contralateral cerebral cortex areas [29]. Notably, the universal cerebellar transform and dysmetria of thought theories propose a cerebellar role in calibrating performance around a homeostatic baseline across motor and non-motor domains – with a domain-general function pertaining to the fine tuning of motor abilities, cognition, behavior, and affect to prevent overshoot and undershoot [30]. These findings and their implications align with a broader literature implicating cerebellar involvement in FND-motor. Prior neuroimaging studies have reported increased cerebellar grey matter volume in FND-motor [31] and in mixed FND patients with more severely impaired mental health [32], heightened cerebellar activation during a motor task in individuals with functional tremor [33], and greater baseline cerebellar functional connectivity predicting subsequent symptom improvement in a mixed FND cohort [34]. Interestingly, a similar pattern of higher FA and NDI, and lower MD, has been reported within the middle cerebellar peduncle and cerebellum in fibromyalgia and somatic symptom disorder [35–37], raising the possibility that alterations within cortico-cerebellar circuitry represent a transdiagnostic feature of disorders with prominent physical symptoms at the brain-mind-body interface.

Comparisons of FND-motor vs. PCs provided additional insight into the specificity of white matter alterations. In contrast to the four-metric convergence observed relative to HCs, FND-motor differed from PCs only in MD and FWF, primarily in posterior corpus callosum, middle cerebellar peduncle, and left inferior longitudinal fasciculus, suggesting that diffusion metrics may differ in their ability to discriminate FND-motor across comparison groups. Robustness of the findings vs. PCs also varied: MD differences were most robust in cerebellar white matter, surviving *post-hoc* adjustments and female-only analyses, whereas FWF differences were most robust in the corpus callosum and inferior longitudinal fasciculus, surviving adjustment for psychiatric symptom burden but attenuating following childhood trauma adjustment and female-only analyses, highlighting the need to consider the role of these factors in future work. The corpus callosum has been implicated in previous diffusion MRI studies of FND, as well as in related conditions such as fibromyalgia [7,8,35,38], however the direction of diffusion abnormalities has varied across studies, warranting further investigation. By contrast, the recurrence of the middle cerebellar peduncle across both HC and PC comparisons identifies this tract as specifically altered in FND-motor.

Within FND-motor, higher FND symptom severity (SDQ-20) was associated with lower FA and NDI, and higher MD and FWF, in the mid-posterior and central corpus callosum and predominantly right-lateralized association and projection pathways. These white matter-SDQ-20 relationships remained significant in *post-hoc* adjustments for affective symptoms and trauma burden, although NDI associations attenuated following adjustment for depression, anxiety, and PTSD severity. This pattern differed from the group-level findings in terms of the implicated pathways and direction of diffusion abnormalities, suggesting that individual differences in white matter alterations associated with FND symptom severity are at least partially distinct from those differentiating FND-motor from HCs and PCs at the group-level. Of relevance, corpus callosum alterations are implicated in disorders of motor agency and ownership – suggesting that disruptions in interhemispheric transmission of information may relate to FND symptom severity [39]. Although not observed as a convergent multi-metric finding, a positive association between FA and SDQ-20 scores was also identified in the middle cerebellar peduncle, and held across *post-hoc* corrections, further highlighting this tract as a recurrent site of white matter differences across analyses.

Across FND-motor and PCs, greater transdiagnostic physical symptom burden (PHQ-15) was associated with higher FA, and lower MD and FWF, in the middle cerebellar peduncle and adjacent cerebellar white matter, a pattern that closely resembled the between-group findings. These associations remained robust following *post-hoc* adjustment for depression, anxiety, and PTSD severity, and were present in female-only analyses, although MD and FWF associations attenuated after adjustment for childhood trauma, suggesting that childhood maltreatment may play a role in some of these associations. Scatterplots revealed that FND-motor and PC participants showed substantial overlap in the low-to-moderate range of PHQ-15 scores, yet FND-motor participants anchored the upper end of the physical symptom burden distribution, suggesting that middle cerebellar peduncle microstructure relates to transdiagnostic physical symptom burden, while also capturing the greater burden characteristic of FND-motor.

Several limitations warrant consideration. FND severity was assessed using a patient-reported measure and should be interpreted as reflecting subjective symptom burden rather than clinician-rated severity. Although the heterogeneous FND-motor cohort enhances generalizability across motor phenotypes, it limits phenotype-specific interpretation. Larger studies examining individual motor phenotypes (including static vs. paroxysmal symptoms), in conjunction with use of clinician-rated and newer patient-rated symptom severity measures, represent an important direction for future work [40]. While the multi-metric approach across DTI and NODDI strengthens confidence through convergent findings, diffusion MRI remains an indirect measure of tissue microstructure [14]. Similarly, probabilistic tractography only estimates likely anatomical pathways, and middle cerebellar peduncle observations are complicated by the dense packing of fibers. Finally, the cross-sectional design precludes causal inferences about observed white matter differences. Future longitudinal studies tracking white matter microstructure over the course of illness and treatment will be important future directions.

In summary, this study provides a comprehensive multi-metric diffusion MRI characterization of white matter microstructure in FND-motor relative to HCs and PCs. The consistent involvement of the middle cerebellar peduncle across group-level comparisons and associations with transdiagnostic physical symptom burden, together with dissociable white matter correlates of FND symptom severity, advances understanding of white matter involvement in FND-motor and provides a foundation for future investigations.

## Data Availability

All data produced in the present study are available upon reasonable request to the authors.

## ACKNOWLEDGMENTS

We thank all the research participants, including those with FND, for their participation.

## DATA AVAILABILITY

For qualified researchers, analysis code and de-identified data pertaining to study results can be made available following local IRB approval upon reasonable request. Requests will be considered after planned analyses and reporting have been completed by the investigators.

## AUTHOR ROLES

1. Research Project: A. Conception, B. Organization, C. Execution; (2) Statistical Analysis: A. Design, B. Execution, C. Review and Critique; (3) Manuscript: A. Writing of the First Draft, B. Review and Critique

CW: 1A, 1B, 1C, 2A, 2B, 3C, 3A, 3B; CB: 1B; 1C; 2C, 3B; AJG: 1B; 1C; 2C, 3B; SAF: 1C, 3B; JM: 1C, 3B; JR: 1C, 3B; JM: 1C, 3B;EG: 1C, 3B; DM: 1C, 3B; JF: 1C, 3B; CA: 1C, 3B; CDS: 1C, 3B; MK: 1A, 3B; ID: 1A, 2C, 3A, 3B; DLP: 1A, 1B, 2A, 2C, 3A, 3B

