## Supplementary Materials for "White Matter Microstructural Alterations and Symptom Correlates in Functional Motor Disorder"

**MRI Acquisition and Preprocessing**

High-resolution T1-weighted magnetization-prepared rapid gradient echo (MP-RAGE) scans were acquired with the following parameters: 1mm isotropic voxels; 160 sagittal slices; acquisition matrix size=256x256; repetition time=2300ms; echo time = 2.98ms; field of view=256mm. Multi-shell diffusion-weighted images were acquired using a spin-echo echo-planar imaging (EPI) sequence with the following parameters: 2mm isotropic voxels; 73 axial slices; acquisition matrix size=128x128; repetition time=10,800ms; echo time=109ms; field of view = 256mm. The acquisition included 8 volumes at b=0 s/mm^2^, 1 volume each at b=50,100,150 s/mm^2^, 3 volumes at b=200 s/mm^2^, 6 volumes at b=500 s/mm^2^, 30 volumes at b=1000 s/mm^2^, and 30 volumes at b=2500 s/mm^2^.

T1-weighted data were preprocessed using FMRIB Software Library v5.0.7 (FSL, Oxford, UK) and MATLAB R2026a (MathWorks, Natick, MA) using in-house preprocessing pipelines (previously described [17]). Anatomical preprocessing included: reorientation to right-posterior-inferior (RPI); denoising and N4 bias-field correction; alignment to the anterior and posterior commissures; skull stripping; segmentation of grey matter, white matter, and cerebrospinal fluid; and nonlinear registration to a 2mm resolution MNI152 template.

Preprocessing of diffusion-weighted images was done using FSL, MRtrix3, and MATLAB using in-house preprocessing pipelines. Preprocessing included: denoising; brain mask generation; and correction for eddy-current distortions and head motion. Average volume-to-volume translation was computed from the affine transformation matrices estimated during eddy-current and motion correction for use as a nuisance covariate in subsequent statistical analyses, reflecting the magnitude of the affine transforms required to align consecutive diffusion-weighted volumes. For conventional diffusion tensor imaging (DTI) metrics, b=0 and b=1000 s/ mm2 shells were extracted followed by local fitting of the diffusion tensor at each voxel to compute fractional anisotropy (FA) and mean diffusivity (MD) maps. For neurite orientation dispersion and density imaging (NODDI), the full multi-shell diffusion data were fit using the NODDI Toolbox in MATLAB, to generate neurite density index (NDI), orientation dispersion index (ODI), and free water fraction (FWF) maps. Boundary-based registration was used to register each diffusion image to each participant’s T1-weighted anatomical image.

**Probabilistic Tractography**

**To characterize the implicated fiber bundles from convergent white matter VBA findings, probabilistic tractography was performed using diffusion-weighted MRI data from the 50 HCs. Binarized cross-metric convergence maps were first transformed from MNI152 standard space into each participant’s native diffusion space. FSL BEDPOSTX was then used to model crossing fibers, after which whole-brain probabilistic tractography was performed using FSL PROBTRACKX2 with 1,000 streamlines initiated from each seed voxel. Individual subject tractography maps were normalized by waytotal, the total number of streamlines initiated from the seed that completed tractography, and were then transformed back to MNI152 standard space. Group-level probabilistic projection maps were then generated by averaging the normalized maps across all HCs, with map values reflecting the mean normalized streamline density traversing each voxel, providing an approximation of the probability of structural connectivity between the seed region and each voxel.**

**Quantifying Tract Overlap**

To anatomically contextualize the white matter findings, two complementary atlas-based tract overlap analyses were performed using binary white matter tract masks from the HCP-1065 Population-Averaged Tractography Atlas [28]. First, to localize the significant white matter findings within established anatomical tracts, the overlap of convergent voxels with each tract was computed. For this, the number of overlapping voxels within each tract was divided by the total number of overlapping voxels across all atlas tracts, expressed as a percent overlap. Second, to characterize the broader white matter pathways most likely to traverse the implicated regions, the overlap of probabilistic tractography maps with each tract was computed. For this, the group-averaged probabilistic tractography maps were thresholded (normalized streamline density > 0.01), and weighted overlap was calculated as the sum of normalized streamline density values within each tract. Percent overlap was computed by dividing each tract's weighted overlap by the summed weighted overlap across all atlas tracts. Whereas direct atlas overlap localized the significant voxels themselves, atlas overlap of the probabilistic tractography maps characterized the broader pathways traversing those voxels. Tracts with percent overlap greater than 5% are reported.

**Supplementary Table 1. Demographic characteristics of participants with functional neurological disorder.**

| **FND Subject** | **Phenotypic Description** | **Current SCID-I**  **Diagnoses** | **Past SCID-I Diagnoses** | **Psychotropic Medications** |
| --- | --- | --- | --- | --- |
| 1 | clinically-established tremor | Specific Phobia, ANX NOS | - | - |
| 2 | documented functional seizures, clinically-established functional tremor & functional speech | AG, Social Phobia, Somatoform Pain Disorder, Undifferentiated Somatoform Disorder | MDE | CLP, AMT |
| 3 | clinically-established functional seizures & clinically-established functional tremor | - | PTSD | - |
| 4 | clinically-established functional gait | ANX NOS, Undifferentiated Somatoform Disorder | DEP NOS, Specific Phobia | - |
| 5 | clinically-established functional limb weakness (left arm/leg) | DYS, MDE, GAD, PD+AG, Somatization Disorder, Hypochondriasis | PTSD | - |
| 6 | documented functional seizures; clinically-established functional limb weakness (left leg) | - | MDE, PTSD, PD-AG | LTG, TZD |
| 7 | clinically-established functional tremor | DYS, PD+AG, PTSD | Eating Disorder, MDE, OCD | LRZ, SERT |
| 8 | clinically-established functional tremor | Somatoform Pain Disorder | ETOH Abuse, ANX NOS | DLX, CLP |
| 9 | clinically-established functional limb weakness (arms/legs) | ANX NOS | MDE, PTSD, Eating Disorder | TZD, ECP, BUP |
| 10 | clinically-established functional tremor & functional gait | MDE, SAD | MDE, PD+AG | FLX, CLP |
| 11 | clinically-established functional tremor & functional speech | Social Phobia, OCD, GAD | DEP NOS | SERT |
| 12 | clinically-established functional limb weakness (left arm/leg) & functional gait | DYS, PD+AG, PTSD, SSD | MDE, Eating Disorder | LDA, CLP, ECP, QTP, BCP |
| 13 | clinically-established functional tremor | GAD, IAD | MDE, PTSD | DLX, LRZ |
| 14 | clinically-established functional jerky movements (head & bilateral upper extremity) | GAD | Eating Disorder, AUD, SUD | ECP, PGB, AMT, APM / DXAM |
| 15 | functional tremor in four-limbs | ANX NOS | - | GBP |
| 16 | clinically-established functional jerky movements | GAD | DEP NOS | SERT |
| 17 | clinically-established functional limb weakness (left arm) & functional speech | GAD, SSD | MDE, PTSD | CBD |
| 18 | clinically-established functional tremor, probable functional seizures | - | MDE | AMT |
| 19 | documented functional seizures, clinically-established functional limb weakness (left leg) & functional gait | MDE, GAD, PD+AG | MDE | CLP, FLX, PZN |
| 20 | clinically-established functional limb weakness (legs) & functional gait | GAD, SSD | MDE | LRZ, BSP, LTG |
| 21 | clinically-established functional tremor & functional speech | PTSD, GAD, PD+AG, MDE | - | DLX, BUP, LRZ |
| 22 | clinically-established functional gait & functional speech | AG, ADHD | - | APZ, CBD |
| 23 | documented functional seizures; clinically-established functional gait & functional speech | ANX NOS | PTSD, PD+AG | PGB, CBD |
| 24 | clinically-established functional gait, functional tremor, functional limb weakness (right arm/leg), & functional speech | GAD, PD+AG | PTSD, MDE | TPM |
| 25 | clinically-established functional gait & functional limb weakness (right arm/leg) | - | MDE, PTSD, Eating Disorder | GBP, AMT, TZD, PZN |
| 26 | clinically-established functional tremor | - | PTSD, BPAD-II (with MDE) | LTM, QTP, LRZ |
| 27 | clinically-established functional limb weakness (legs) & functional speech | - | DEP NOS, Specific Phobia | - |
| 28 | clinically-established functional tremor, functional dystonia & functional gait | PD+AG, SSD | GAD, DEP NOS | - |
| 29 | clinically-established functional facial spasms/tics & functional speech | GAD, PD+AG, ADHD, PTSD | MDE | CLN, LDA, APR, SERT, LRZ |
| 30 | clinically-established functional gait & functional speech | - | DEP NOS, ANX NOS | GBP |
| 31 | clinically-established functional limb weakness (left leg) | ADHD | DYS, MDE, GAD, SAD | DLX, LDA, PGB |
| 32 | clinically-established functional jerks/spasms/tics | PTSD, ADHD | SAD, MDE, Specific Phobia | MIR, SERT, MPD, LRZ |
| 33 | clinically-established functional tremor (right arm & leg) & functional dystonia (right foot) | MDE, GAD, PTSD | MDE, PD+AG, SAD | BSP |
| 34 | clinically-established functional limb weakness (left leg) | ANX NOS | MDE, ANX NOS | GBP, HDZ, ECP |
| 35 | probable functional seizures; clinically-established functional limb weakness (bilateral leg), functional tremor, & functional gait | SSD, AG, DYS | MDE, PD-AG, GAD, PTSD, Eating Disorder | AMT, DLX |
| 36 | clinically-established functional limb weakness (4-limb) & functional tremor | SSD, SAD, GAD, PD | PTSD, ADHD | SERT, CLP |
| 37 | clinically-established functional dystonia & functional speech | GAD | PTSD | - |
| 38 | clinically-established functional limb weakness (left arm & leg), functional tremor, & functional jerks | - | PTSD | - |
| 39 | clinically-established functional tremor (right arm & leg) | PTSD, MDE, Anxiety NOS | - | MIR, CLP |
| 40 | clinically-established functional tremor* | PTSD, MDE, SAD, GAD, SSD | DYS, PTSD, AUD, ANX NOS | APZ, BSP; PZN, ECP, TZD, HDZ |
| 41 | clinically-established functional movement disorder (functional limb weakness (arms and left leg), functional jerks) | PD+AG, MDD, SAD, GAD, SSD, IAD, ADHD | - | QTP, LTG, APM, HDZ |
| 42 | documented functional seizures; clinically-established* functional jerks/tics, & functional speech | PD+AG, DYS, GAD, SSD | OCD | DLX, GBP, LRZ |
| 43 | clinically-established functional tremor, & functional gait | Anxiety NOS | - | HDZ |
| 44 | documented functional seizures; clinically-established functional gait, functional limb weakness (left arm and leg), & functional speech | PTSD, MDE, PD + AG, GAD, SSD | MDE, Eating Disorder, AUD, SUD | BUP, GBP, TZD, HDZ |
| 45 | clinically-established functional movement disorder (right functional leg weakness, bilateral functional tremor in arms, functional gait) | - | PTSD, MDE | - |
| 46 | clinically-established functional movement disorder (functional weakness (4-limb), functional tremor (4-limb), functional jerks, functional gait) | SSD | - | - |
| 47 | clinically-established functional movement disorder (functional gait, functional weakness (left leg), functional tremor (hands), and functional speech) | Specific Phobia | MDD | - |
| 48 | clinically-established functional limb weakness (left arm and leg) & functional gait | - | MDE, SAD, PTSD, Eating Disorder | CTP |
| 49 | clinically established functional movement disorder (gait, weakness) | ANX NOS | PD-AG | ECP |
| 50 | documented functional seizures; clinically-established functional movement disorder (gait, dystonic posturing in bilateral feet) | MDE, GAD | MDE, Eating Disorder | BUP, GBP, CLP |

*Indicates subject also had concurrent functional somatosensory loss (e.g., non-dermatomal somatosensory deficits). Subjects 1-11 were evaluated using a SCID-I for DSM-IV-TR, while subjects 12-50 were evaluated using the SCID-I for DSM-5; subject 45 had missing SCID-I data and psychiatric comorbidities are based on chart-review diagnoses. ADHD, Attention-Deficit/Hyperactivity Disorder; AG, Agoraphobia; ANX, Anxiety; AUD, Alcohol Use Disorder; BPAD, Bipolar Affective Disorder; DEP, Depression; DYS, Dysthymia; ETOH-Abuse, Alcohol Abuse; GAD, Generalized Anxiety Disorder; IAD, Illness Anxiety Disorder; MDE, Major Depressive Episode; NOS, not otherwise specified; OCD, Obsessive Compulsive Disorder; PD+AG, Panic Disorder with Agoraphobia; PD-AG, Panic Disorder without Agoraphobia; PTSD, Post-Traumatic Stress Disorder; SAD, Social Anxiety Disorder; SSD, Somatic Symptom Disorder; SUD, Substance Use Disorder; AMT, Amitriptyline; APM, Amphetamine; APR, Aripiprazole; APZ, Alprazolam; BCP, Baclophen; BSP, Buspirone; BTP, Benztropine; BUP, Bupropion; CBD, Cannabidiol; CLN, Clonidine; CLP, Clonazepam; CTP, Citalopram; DLX, Duloxetine; DVX, Desvenlafaxine; DXAM, Dextroamphetamine; DZP, Diazepam; ECP, Escitalopram; FLX, Fluoxetine; GBP, Gabapentin; HDZ, Hydroxyzine; LDA, Lisdexamfetamine; LTM, Lithium; LTG, Lamotrigine; LRZ, Lorazepam; LTA, Levetiracetam; MIR, Mirtazapine; MLT, Melatonin; MPD, Methylphenidate; NRT, Nortriptyline; OLZ, Olanzapine; PGB, Pregabalin; PZN, Prazosin; QTP, Quetiapine; SERT, Sertraline; TPM, Topiramate; TZD, Trazodone.

**Supplementary Table 2. Demographic characteristics of psychiatric controls with a lifetime history of clinically-salient depression, anxiety and/or post-traumatic stress disorder.**

| **PC**  **Subject** | **Current SCID-I**  **Diagnoses** | **Past SCID-I**  **Diagnoses** | **Psychotropic Medications** |
| --- | --- | --- | --- |
| 1 | DEP NOS, PD+AG, PTSD, Social Phobia | MDE | DLX, CLP, LTG, QTP |
| 2 | MDE | MDE, ANX NOS | BUP, DLX |
| 3 | MDE, PTSD | MDE, AUD | ZPD, MPD, TZD, MIR, GBP, TPM |
| 4 | MDE, GAD | PTSD | BUP, LTG, BSP |
| 5 | - | MDE, ANX NOS | - |
| 6 | ANX NOS | MDE | - |
| 7 | - | MDE | - |
| 8 | - | PTSD, MDE | - |
| 9 | - | DEP NOS | - |
| 10 | - | PTSD | - |
| 11 | - | MDE | - |
| 12 | GAD | MDE, Eating Disorder, Anxiety NOS | FLX, LRZ |
| 13 | MDE, GAD | PTSD | FLX, MIR |
| 14 | MDE, DYS, GAD, AG | - | ECP, APR |
| 15 | - | ANX NOS, DEP NOS | VEN, BUP |
| 16 | DEP NOS, ANX NOS | MDE | SERT, QTP, GBP |
| 17 | GAD | MDE | BUP, MLT |
| 18 | DYS, ANX NOS | MDE, AUD | BSP, SERT |
| 19 | GAD, SAD | - | SERT |
| 20 | MDE, PTSD, GAD, OCD | - | BSP, LTG, BUP, APM / DXAM, TZD, MLT |
| 21 | Eating Disorder | MDE, PD-AG, OCD | - |
| 22 | ANX NOS | DEP NOS, GAD | SERT |
| 23 | - | GAD, DEP NOS | BUP |
| 24 | - | ANX NOS | SERT |
| 25 | - | MDE, Specific Phobia | - |
| 26 | - | MDE, PD+AG | - |
| 27 | GAD | MDE | - |
| 28 | - | MDE, PTSD, PD+AG, OCD, AUD | - |
| 29 | PTSD, SAD, ADHD, PD+AG | MDE, Eating Disorder | LDA |
| 30 | - | MDE | GBP, CTP |
| 31 | GAD, MDE, SAD, Specific Phobia | DYS, PTSD | CLP |
| 32 | PTSD | MDE | ECP, CLN |
| 33 | GAD, MDE | - | ECP |
| 34 | MDE, Anxiety NOS, MDE | GAD, MDE, AUD, PD+AG | - |
| 35 | PTSD, GAD, ADHD | AUD | LRZ |
| 36 | PTSD, GAD, MDE, DYS | Eating Disorder | VEN, MPD, LRZ, PZN, QTP, GBP, BPN, EZP |
| 37 | PTSD, OCD, SAD | - | FLX |
| 38 | PTSD, MDD, GAD | - | SERT, ECP, TZD, APZ |
| 39 | - | MDD | BUP, APM, TZD, VEN |
| 40 | MDD, GAD | - | LDA, VEN, MIR |
| 41 | MDD, SAD | PTSD, SUD | CTP, MPD |
| 42 | MDD, PTSD | PD+AG | DLX, APM, TZD |
| 43 | - | MDD, PTSD | - |
| 44 | PTSD, MDD, IAD | - | - |
| 45 | SAD | PTSD, MDD | BUP, MPD, ECP |
| 46 | PD - AG, OCD | Eating Disorder NOS, MDD, SAD | LDA, ECP |
| 47 | - | GAD | FLX, LDA |
| 48 | GAD, Specific Phobia, ADHD | MDD | ATM |
| 49 | DEP NOS, ANX NOS, Specific Phobia | MDD, AG | VEN, BUP, APM / DXAM |
| 50 | SAD | - | - |

Subjects 1-4 were evaluated using a SCID-I for DSM-IV-TR, while subjects 5-50 were evaluated using the SCID-I for DSM-5. Clinically-salient depression was defined as any subject with diagnoses of MDE (including in the context of BPAD), DEP NOS, DYS, and/or MDD. Clinically-salient anxiety was defined as AG, ANX NOS, GAD, OCD, PD ± AG, SAD, IAD, Specific Phobia, and/or Social Phobia. ADHD, Attention-Deficit/Hyperactivity Disorder; AG, Agoraphobia; ANX, Anxiety; AUD, Alcohol Use Disorder; BPAD, Bipolar Affective Disorder; DEP, Depression; DYS, Dysthymia; ETOH-Abuse, Alcohol Abuse; GAD, Generalized Anxiety Disorder; IAD, Illness Anxiety Disorder; MDD, Major Depressive Disorder; MDE, Major Depressive Episode; NOS, not otherwise specified; OCD, Obsessive Compulsive Disorder; PD+AG, Panic Disorder with Agoraphobia; PD-AG, Panic Disorder without Agoraphobia; PTSD, Post-Traumatic Stress Disorder; SAD, Social Anxiety Disorder; APM / DXAM, Amphetamine / Dextroamphetamine; APR, Aripiprazole; APZ, Alprazolam; ATM, Atomoxetine; BPN, Buprenorphine-naloxone; BSP, Buspirone; BUP, Bupropion; CLN, Clonidine; CLP, Clonazepam; CTP, Citalopram; DLX, Duloxetine; ECP, Escitalopram; EZP, Eszopiclone; FLX, Fluoxetine; GBP, Gabapentin; LDA, Lisdexamfetamine; LRZ, Lorazepam; LTG, Lamotrigine; LTM, Lithium; MIR, Mirtazapine; MLT, Melatonin; MPD, Methylphenidate; PAX, Paroxetine; PZN, Prazosin; QTP, Quetiapine; SERT, Sertraline; TPM, Topiramate; TZD, Trazodone; VEN, Venlafaxine; ZPD, Ziprasidone.

**Supplementary Table 3.** Overlap between probabilistic tractography projections seeded from convergent between-group voxel-based analysis clusters and HCP-1065 white matter tracts.

| **Between-Group Voxel-Based Analyses** | | | | |
| --- | --- | --- | --- | --- |
|  | **Overlapping HCP-1065 Atlas Tract** | **Weighted Overlap** | **Number of Overlapping Voxels** | **Percent of Total Weighted Overlap** |
| **FND-motor vs. HC**  VBA Convergent Voxels Across FA, MD, NDI, FWF | Middle Cerebellar Peduncle | 32.6 | 1033 | 73.2% |
|  | Left Corticospinal Tract | 3.6 | 249 | 8.1% |
|  | Left Parietal Corticopontine Tract | 3.0 | 181 | 6.7% |
| **FND-motor vs. PC**  VBA Convergent Voxels Across MD, FWF | Corpus Callosum | 22.9 | 1687 | 31.3% |
|  | Left Inferior Longitudinal Fasciculus | 11.9 | 885 | 16.3% |
|  | Anterior Commissure | 11.6 | 777 | 15.9% |
|  | Left Inferior Fronto-Occipital Fasciculus | 8.9 | 616 | 12.2% |
|  | Left Optic Radiation | 8.7 | 582 | 11.9% |
| **Associations With Clinical Features** | | | | |
|  | **Overlapping HCP-1065 Atlas Tract** | **Weighted Overlap** | **Number of Overlapping Voxels** | **Percent of Total Weighted Overlap** |
| **Associations with SDQ-20**  VBA Convergent Voxels Across FA, MD, NDI | Corpus Callosum | 256.6 | 9979 | 26.7% |
|  | Right Parietal Corticopontine Tract | 195.9 | 6298 | 20.4% |
|  | Right Superior Thalamic Radiation | 141.1 | 4483 | 14.7% |
|  | Right Medial Lemniscus | 116.0 | 3555 | 12.1% |
|  | Right Superior Corticostriatal Tract | 112.6 | 3589 | 11.7% |
| **Associations with SDQ-20**  VBA Convergent Voxels Across FA, MD, FWF | Right Parietal Corticopontine Tract | 57.3 | 3446 | 13.2% |
|  | Corpus Callosum | 46.2 | 2830 | 10.7% |
|  | Right Superior Thalamic Radiation | 40.9 | 2534 | 9.5% |
|  | Right Superior Corticostriatal Tract | 40.9 | 2634 | 9.4% |
|  | Right Frontal Corticopontine Tract | 38.7 | 2499 | 8.9% |
|  | Right Superior Corticostriatal Tract | 34.6 | 2200 | 8.0% |
|  | Right Superior Longitudinal Fasciculus 2 | 28.7 | 1717 | 6.6% |
|  | Right Dentatorubrothalamic Tract | 26.9 | 1728 | 6.2% |
|  | Right Medial Lemniscus | 21.7 | 1329 | 5.0% |
| **Associations with PHQ-15**  VBA Convergent Voxels Across FA, MD, FWF | Middle Cerebellar Peduncle | 70.8 | 1896 | 76.3% |
|  | Left Cerebellum WM | 12.5 | 734 | 13.4% |

Probabilistic tractography maps were thresholded at 0.01 prior to overlap calculations. Overlapping voxels indicate the number of suprathreshold projection voxels located within each tract ROI. Weighted overlap represents the sum of projection values across all overlapping voxels, reflecting both the spatial extent and strength of the projection within a given tract. Percent of total weighted overlap indicates the proportion of total weighted overlap across all HCP-1065 trat ROIs accounted for by each tract. Results are shown for tracts with >5 percent of total weighted overlap. HCs, healthy controls; PC, psychiatric controls; VBA, voxel-based analysis; FA, fractional anisotropy; MD, mean diffusivity; NDI, neurite density index; FWF, free water fraction; WM, white matter.

**Supplementary Figure Captions**

**Supplementary Figure 1. Distribution of functional motor phenotypes across the cohort.** An UpSet plot is used to visualize the distribution and co-occurrence of functional motor phenotypes across participants. Vertical bars reflect the number of individuals with each combination of co-occurring motor phenotypes, as shown by multiple darkened circles connected by lines, or a single motor phenotype indicated by one darkened circle. Horizontal bars reflect the total number of participants exhibiting each individual motor phenotype (with or without other co-occurring motor phenotypes).

**Supplementary Figure 2. Voxel-based white matter microstructural alterations in functional motor disorder (FND-motor) compared with healthy controls (HCs) across primary, *post-hoc,* and female-only analyses.** Results from voxel-based analyses comparing individuals with FND-motor (N=50) and HCs (N=50) across fractional anisotropy (FA), mean diffusivity (MD), neurite density index (NDI), free water fraction (FWF), and orientation dispersion index. The top row depicts results from the primary adjustment for age, sex, head motion, SSRI/SNRI use, and co-occurring functional seizures. The second row depicts results from the *post-hoc* adjustment controlling additionally for BDI-II, STAI-total, and PCL-5 scores. The third row depicts results from the *post-hoc* adjustment controlling additionally for CTQ-abuse and CTQ-neglect scores. The bottom row depicts female-only analyses using the primary adjustment. Colors reflect the z-statistic computed from a voxel-wise general linear model (z>1.96, p<0.05, cluster-corrected for multiple comparisons). Warm colors indicate higher diffusion metric values in FND-motor relative to HCs, whereas cool colors indicate lower values. SSRI/SNRI, selective serotonin reuptake inhibitor/serotonin-norepinephrine reuptake inhibitor; BDI-II, Beck Depression Inventory-II; STAI-total, State-Trait Anxiety Inventory-total; PCL-5, PTSD Checklist for DSM-5; CTQ, Childhood Trauma Questionnaire.

**Supplementary Figure 3. Voxel-based white matter microstructural alterations in functional motor disorder (FND-motor) compared with psychiatric controls (PCs) across primary, *post-hoc,* and female-only analyses.** Results from voxel-based analyses comparing individuals with FND-motor (N=50) and PCs (N=50) across mean diffusivity (MD) and free water fraction (FWF). The top row depicts results from the primary adjustment for age, sex, head motion, SSRI/SNRI use, and co-occurring functional seizures. The second row depicts results from the *post-hoc* adjustment controlling additionally for BDI-II, STAI-total, and PCL-5 scores. The third row depicts results from the *post-hoc* adjustment controlling additionally for CTQ-abuse and CTQ-neglect scores. The bottom row depicts female-only analyses using the primary adjustment. Colors reflect the z-statistic computed from a voxel-wise general linear model (z>1.96, p<0.05, cluster-corrected for multiple comparisons). Warm colors indicate higher diffusion metric values in FND-motor relative to PCs, whereas cool colors indicate lower values. SSRI/SNRI, selective serotonin reuptake inhibitor/serotonin-norepinephrine reuptake inhibitor; BDI-II, Beck Depression Inventory-II; STAI-total, State-Trait Anxiety Inventory-total; PCL-5, PTSD Checklist for DSM-5; CTQ, Childhood Trauma Questionnaire.

**Supplementary Figure 4. Diffusion metrics plotted by functional motor disorder (FND-motor) phenotype. (A)** Mean diffusion values extracted from the convergent voxels identified in the FND-motor vs. healthy control (HC) comparison are shown separately for the overall FND-motor cohort (N=50), and by individual motor phenotype. Plots are shown for fractional anisotropy (FA), mean diffusivity (MD), neurite density index (NDI), and free water fraction (FWF). **(B)** Mean diffusion values extracted from the convergent voxels identified in the FND-motor versus psychiatric control (PC) comparison are shown for the overall FND-motor cohort (N=50) and for individual motor phenotype. Plots are shown for mean diffusivity (MD) and free water fraction (FWF). Individual dots represent the diffusion value for a given participant. Counts of each phenotype are as follows: tremor=24, weakness=22; gait=20; tics/jerks/spasms=8; dystonia=4; motor phenotypes were not mutually exclusive.

**Supplementary Figure 5. Whole-brain white matter associations with SDQ-20 symptom severity across primary, *post-hoc,* and female-only analyses.** Results from voxel-wise analyses relating DSDQ-20 scores to fractional anisotropy (FA), mean diffusivity (MD), neurite density index (NDI), and free water fraction (FWF) across individuals with functional motor disorder (FND-motor; N=50). The top row depicts results from the primary adjustment for age, sex, head motion, SSRI/SNRI use, and co-occurring functional seizures. The second row depicts results from the *post-hoc* adjustment controlling additionally for BDI-II, STAI-total, and PCL-5 scores. The third row depicts results from the *post-hoc* adjustment controlling additionally for CTQ-abuse and CTQ-neglect scores. The bottom row depicts female-only analyses using the primary adjustment. Colors reflect the z-statistic computed from a voxel-wise general linear model (z>1.96, p<0.05, cluster-corrected for multiple comparisons). Warm colors indicate positive associations with SDQ-20 scores, whereas cool colors indicate negative associations. SDQ-20, Somatoform Dissociation Questionnaire-20; SSRI/SNRI, selective serotonin reuptake inhibitor/serotonin-norepinephrine reuptake inhibitor; BDI-II, Beck Depression Inventory-II; STAI-total, State-Trait Anxiety Inventory-total; PCL-5, PTSD Checklist for DSM-5; CTQ, Childhood Trauma Questionnaire.

**Supplementary Figure 6. Whole-brain white matter associations with PHQ-15 symptom severity across primary, *post-hoc,* and female-only analyses.** Results from voxel-wise analyses relating PHQ-15 scores to fractional anisotropy (FA), mean diffusivity (MD), neurite density index (NDI), free water fraction (FWF), and orientation dispersion index (ODI) across individuals with functional motor disorder (FND-motor; N=50) and psychiatric controls (PCs; N=50). The top row depicts results from the primary adjustment for age, sex, head motion, SSRI/SNRI use, and co-occurring functional seizures. The second row depicts results from the *post-hoc* adjustment controlling additionally for BDI-II, STAI-total, and PCL-5 scores. The third row depicts results from the *post-hoc* adjustment controlling additionally for CTQ-abuse and CTQ-neglect scores. The bottom row depicts female-only analyses using the primary adjustment. Colors reflect the z-statistic computed from a voxel-wise general linear model (z>1.96, p<0.05, cluster-corrected for multiple comparisons). Warm colors indicate positive associations with PHQ-15 scores, whereas cool colors indicate negative associations. PHQ-15, Patient Health Quesitonnaire-15; SSRI/SNRI, selective serotonin reuptake inhibitor/serotonin-norepinephrine reuptake inhibitor ; BDI-II, Beck Depression Inventory-II; STAI-total, State-Trait Anxiety Inventory-total; PCL-5, PTSD Checklist for DSM-5; CTQ, Childhood Trauma Questionnaire.

**Supplementary Figure 7. Implicated white matter tracts associated with symptom severity. (A)** Tractography seeded from the convergent voxels identified in the SDQ-20 analyses across FA, MD, and neurite density index (NDI). **(B)** Group-averaged tractography seeded from the convergent voxels identified in the SDQ-20 analyses across FA, MD, and FWF. **(C)** Probabilistic tractography seeded from the convergent voxels identified in the PHQ-15 analyses (convergence across fractional anisotropy [FA], mean diffusivity [MD], and free water fraction [FWF]). Tractography was performed across the healthy control (HC) cohort (N=50). indicate the relative density of probabilistic streamlines traversing each voxel, averaged across HCs, with brighter yellow indicating greater streamline density. Maps were thresholded at a normalized streamline density >0.01 for visualization. Corresponding atlas-based tract overlap analyses identifying the implicated white matter tracts are summarized in **Supplementary Table 3**.
