## Supplementary figures and images for "White Matter Microstructural Alterations and Symptom Correlates in Functional Motor Disorder"

### Supplementary Figure 1

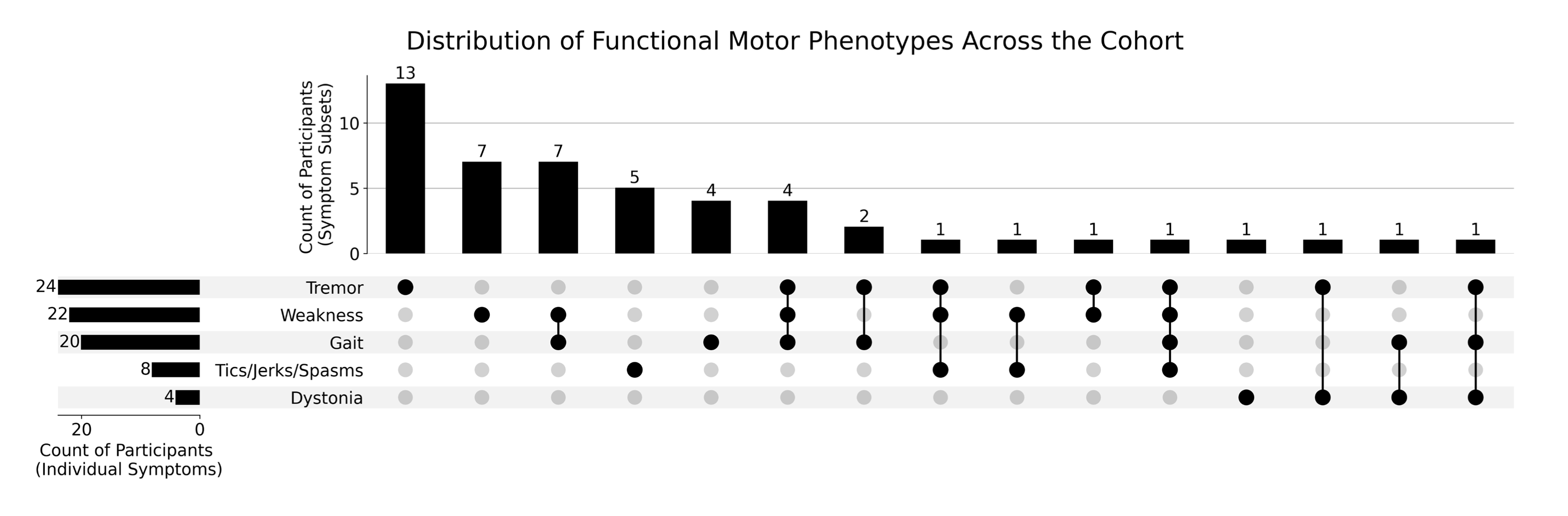

### Supplementary Figure 2

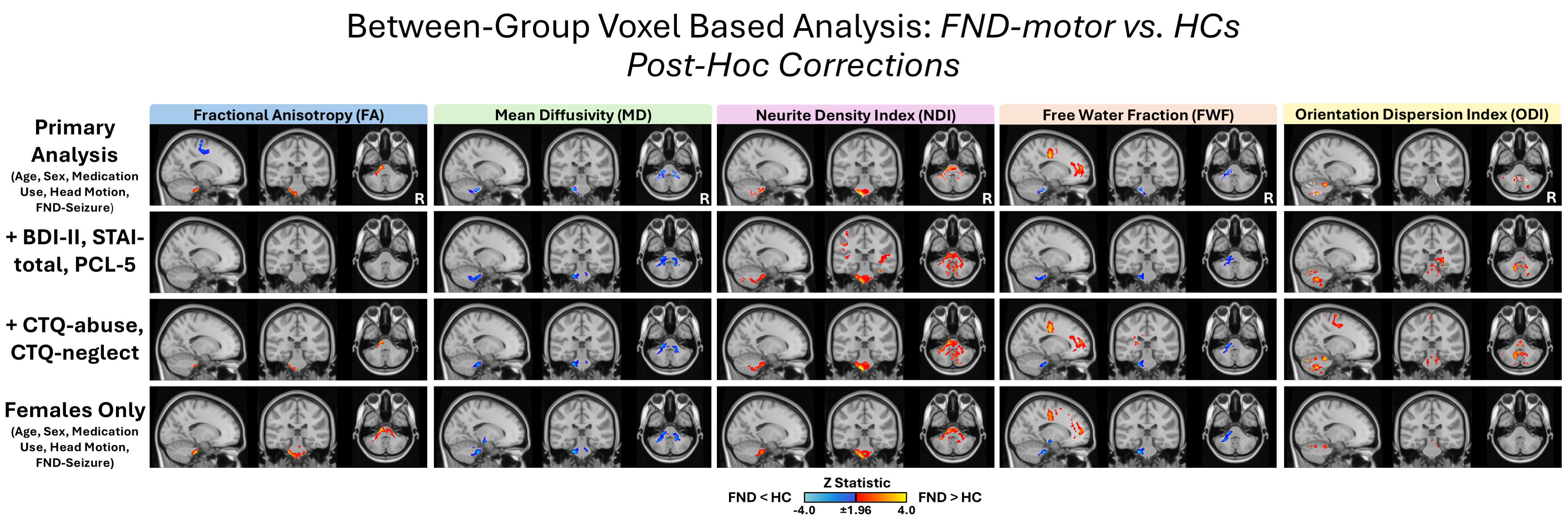

### Supplementary Figure 3

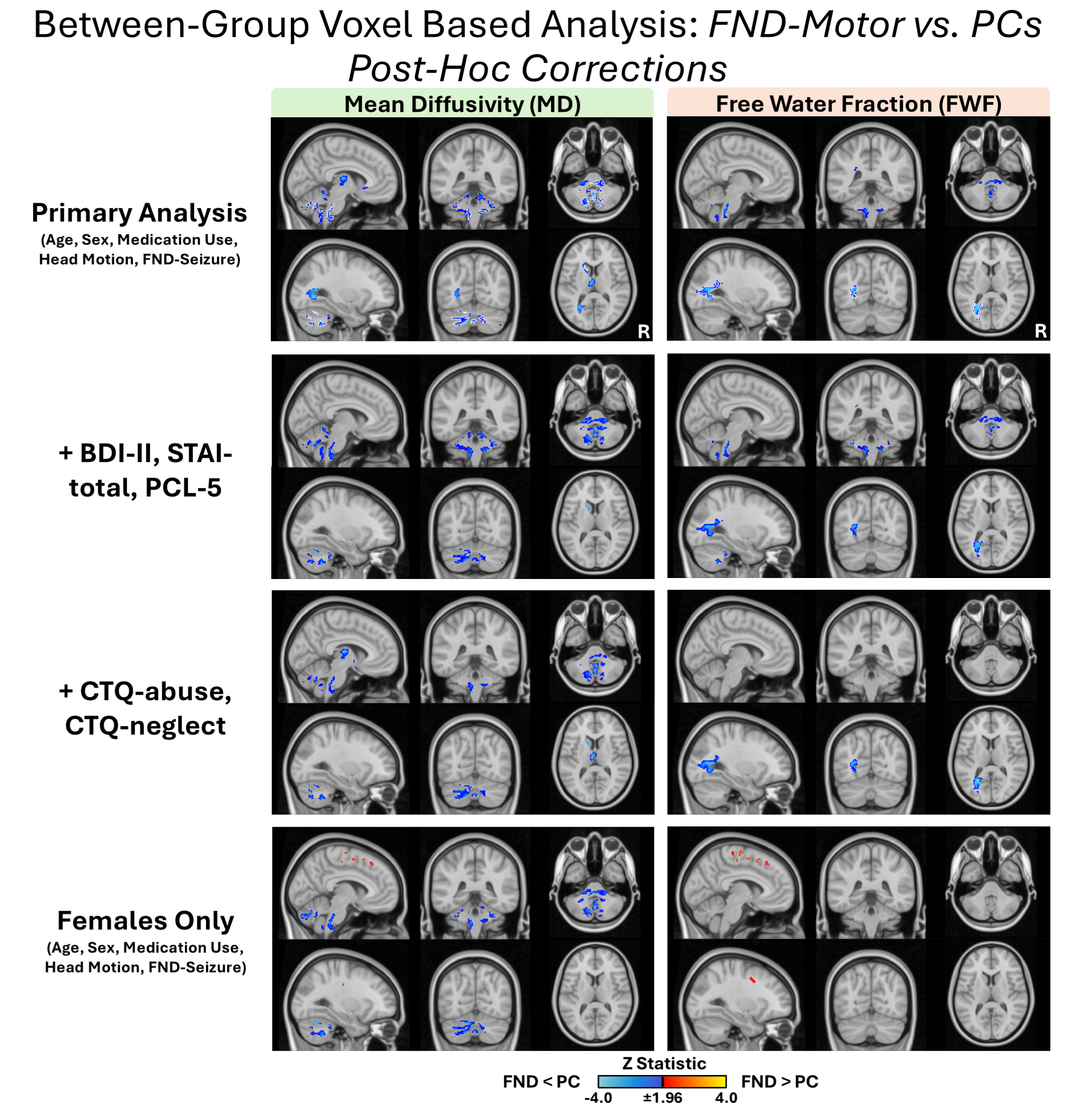

### Supplementary Figure 4

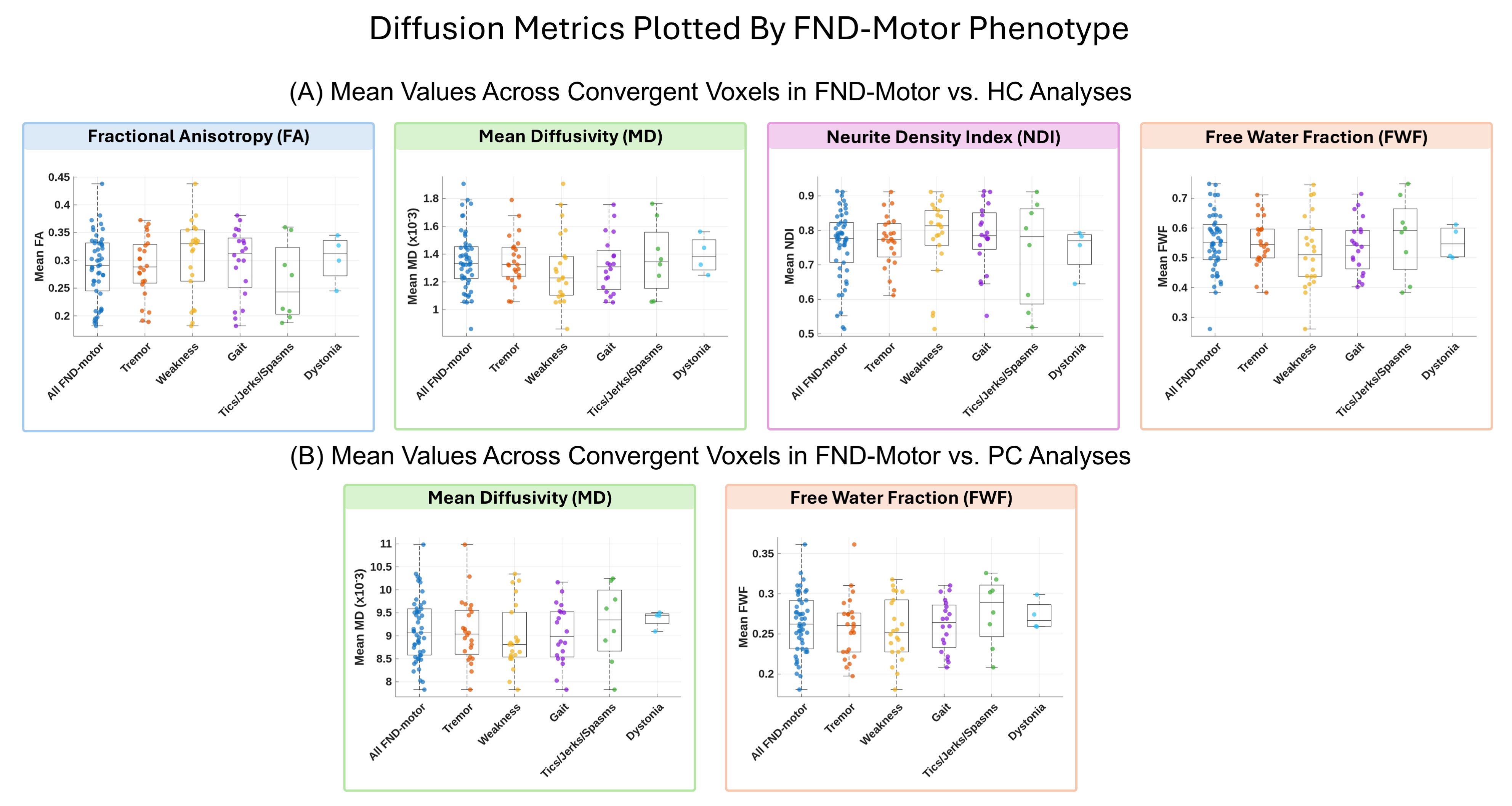

### Supplementary Figure 5

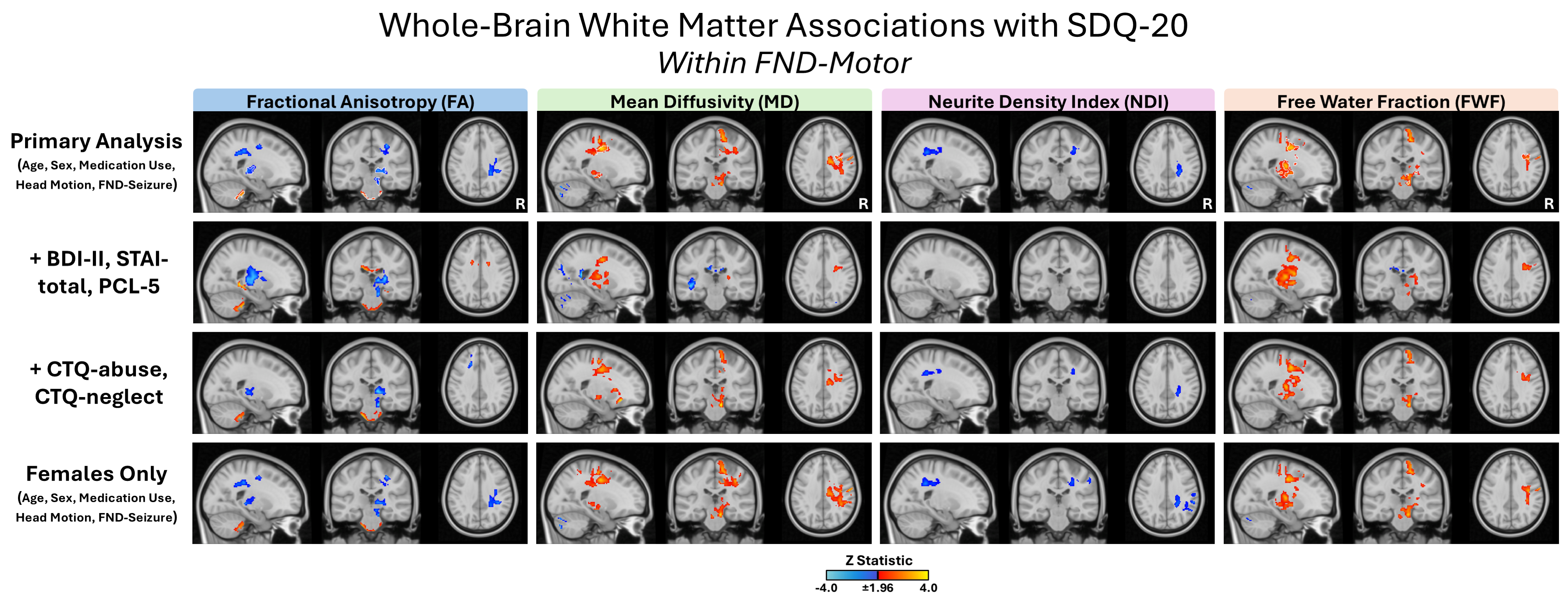

### Supplementary Figure 6

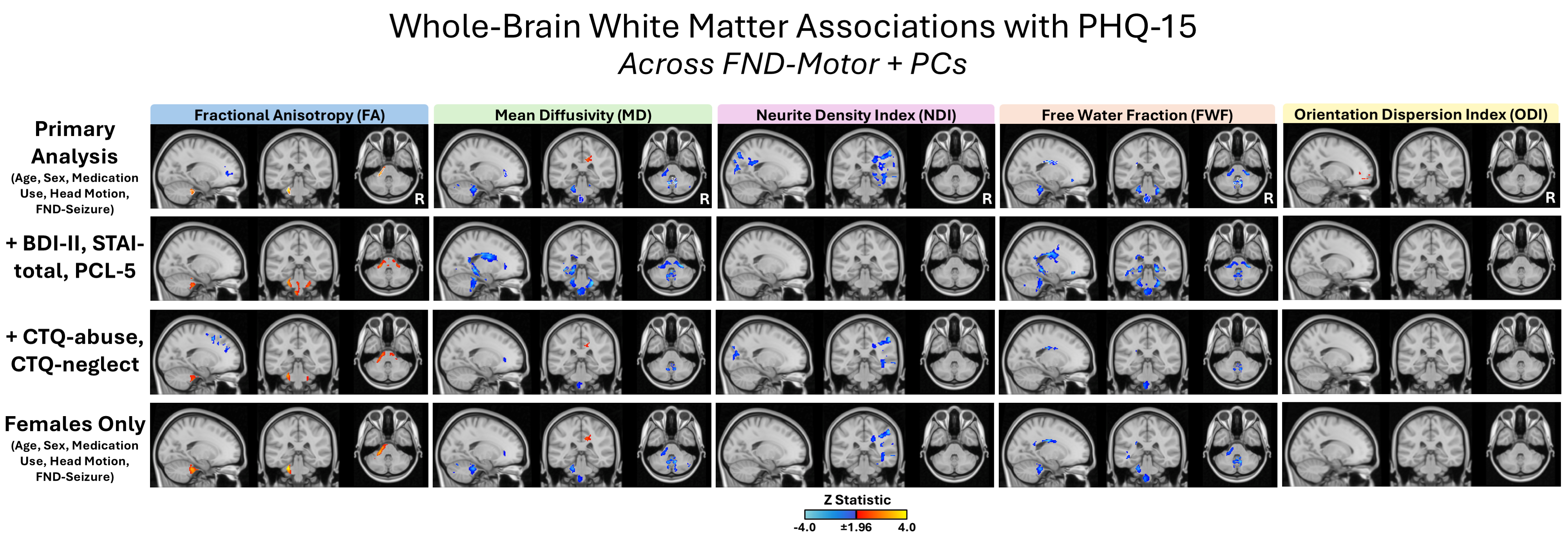

### Supplementary Figure 7

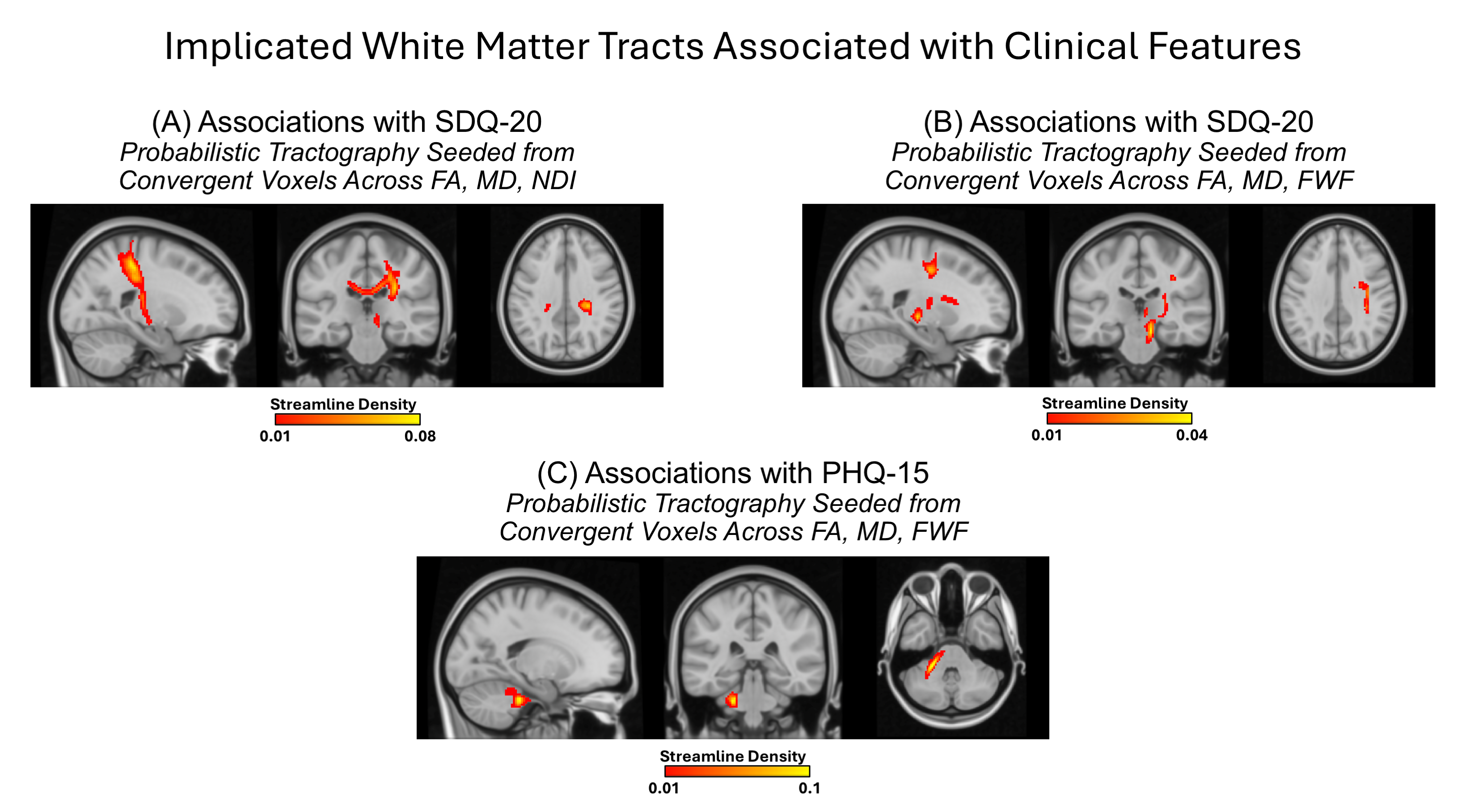
